# Continuous Positive Airway Pressure (CPAP) for Moderate to Severe Covid19 Acute Respiratory Distress Syndrome (CARDS) in a Resource limited setting

**DOI:** 10.1101/2021.06.17.21258809

**Authors:** Anbesan Hoole, Sahar Qamar, Ayesha Khan, Mariam Ejaz

**Affiliations:** Department of Internal Medicine, Bach Christian Hospital, Karakoram Highway, Qalandarabad, Abbottabad, KPK, Pakistan. 22000; Department of Internal Medicine, Bach Christian Hospital, Qalandarabad, Abbottabad, KPK, Pakistan

**Keywords:** ARDS, COVID-19, Non invasive ventilation, Critical Care, Pneumonia

## Abstract

**Introduction:** Covid19 Acute Respiratory Distress Syndrome (CARDS) poses a challenge in management particularly due to limited capacity of ventilated intensive care beds and staffing, and this is exacerbated in resource poor settings with poor patient outcomes. Within this context CPAP has been trialled for CARDS although mainly in resource rich settings.

**Methods:** This study retrospectively analyses the survival outcomes and characteristics of a cohort of patients with moderate to severe CARDS were treated exclusively with CPAP in a rural secondary level hospital in Pakistan with limited previous critical care expertise.

**Results:** 32 out of the 41 patients (78%) who were treated with CPAP survived overall (30/37 (81%) who were treated according to protocol).

**Discussion:** Results suggest non inferiority to CARDS outcomes of critical care units employing Intubation and Mechanical Ventilation (IMV) in resource rich settings. CPAP should be promoted as an efficacious and cost-effective method for treating CARDS within the context of the pandemic surge of Covid19 in resource poor settings.

**Key Messages:** *What is the key question?:* Is Continuous Positive Airway Pressure (CPAP) an effective treatment for Covid19 Acute Respiratory Distress Syndrome (CARDS) in a resource poor setting in a pandemic surge context?

*What is the bottom line?:* Survival rate for CARDS on CPAP in our single centre retrospective cohort study is 78% which is similar to outcomes from critical care centres in resource rich settings employing Intubation and Mechanical Ventilation (IMV) and better than outcomes in many critical care centres in resource poor settings. This suggests CPAP should be promoted as an efficacious and cost-effective method for managing the pandemic surge of CARDS in resource poor settings.

*Why read on?:* The current surge of Covid19 CARDS in resource poor settings poses a significant challenge in terms of effective management given cost and resource restraints, reflected by poor outcomes in overwhelmed critical care centres employing IMV. This is the largest study so far documenting the survival outcomes and characteristics of patients with CARDS treated exclusively with CPAP within a resource poor setting.

## Introduction

Severe Acute Respiratory Syndrome Coronavirus 2 (SARS-CoV-2) causes a Pneumonia requiring hospital admission in 15% of patients(1), of which around a third develop Covid19 Acute Respiratory Distress Syndrome (CARDS) with a mortality rate ranging between 30 – 60%(2). CARDS is defined as Acute Hypoxemic Respiratory failure occurring within 1 week of onset of respiratory symptoms, with bilateral opacities on chest X ray or CT Chest, not explained by fluid overload or heart failure and a SpO_2_/FiO_2_ < 315(3). Intubation and Mechanical Ventilation (IMV) was initially recommended by the WHO for patients with moderate to severe ARDS (P_a_O_2_/FiO_2_ < 150mmHg)(3). However even developed countries have struggled to provide the ventilator and nursing capacity for the surge of Covid19 ARDS patients(4), and the critical care capacity in low- and middle-income countries (LMIC) is much less(5), and this has been reflected in dire outcomes for CARDS(6). For example, a recent review of critical care outcomes for CARDS patients in Africa revealed that around only 50% of patients referred to critical care being admitted, and only 50% of admitted patients surviving(7). Within this context, CPAP is a much less resource intensive option than IMV and was trialled for CARDS patients in Italy early in the pandemic with promising results(8). Studies conducted in various settings since then have produced mixed results on the effectiveness of CPAP in CARDS compared with IMV(9–15), but CPAP still retains its place in prominent guidelines(16,17). However, studies from resource limited settings, which now account for the majority of new Covid19 cases(18) and deaths have been limited. This paper retrospectively analyses the survival outcomes and characteristics of exclusive use of CPAP for moderate to severe CARDS in a cohort of patients at a secondary level hospital in rural Northern Pakistan (Bach Christian Hospital, Khyber Pakhtunkhwa)

## Methods

### Inclusion criteria

Data was collected retrospectively of patients admitted to the Covid19 unit at Bach Christian Hospital (BCH) between April and May 2021 during the 3 ^rd^ wave of Covid19 in Pakistan. Patients were admitted in Covid19 unit based on Respiratory failure (SpO2 < 90% or Respiratory Rate > 30), in the presence of a clinical history and examination suggestive of Covid19 pneumonia (Acute onset fever, cough, dyspnea, crepitations on lung auscultation) with appropriate radiological and laboratory findings (Infiltrates on CXR +/- lymphopenia on CBC)(14). SARS-CoV-2 PCR was performed on affording patients but limited due to cost and availability(20). However, all patients tested had positive results. All patients who presented with or developed moderate to severe ARDS (acute onset respiratory failure with bilateral infiltrates on CXR, not explained by heart failure or fluid overload, P_a_O_2_/FiO_2_ < 150mmHg) were commenced on CPAP and included in this cohort being followed until discharge.

Ethical approval was waived due to the observational nature of this study.

### Management Protocol

A Covid19 unit was opened at BCH in December 2020 to respond to the surge of severely unwell Covid19 patients in Pakistan’s second wave. A 9 bedded unit with isolated beds including 5 with continuous monitoring capacity was allocated for Covid19 patients. Nursing staff with limited previous critical care experience were trained in the management of critically unwell patients, the use of CPAP and Arterial Blood Gas (ABG) sampling. The unit was staffed by a single nurse and nurse aid. CPAP was chosen over High Flow Nasal Cannula (HFNC) for patients with CARDS to conserve oxygen supply which was sourced from an on-site generator plant.

### Respiratory Support

#### Initial Management and CPAP initiation

Patients with acute hypoxemic respiratory failure (AHRF) and clinical findings suggestive of Covid19 pneumonia were initially resuscitated with 5L O_2_ via Nasal Cannula (NC)(21) or 15L O2 via Non-Rebreathe Mask (NRM) depending on severity with a target SpO2 of 95% (fig 1). After 1h clinical assessment with an ABG was performed and those with persistent tachypnoea or P_a_O_2_/FiO_2_ < 150mmHg (corresponding to requiring more than 5L O2 via nasal cannula to maintain SpO2 > 90%) were commenced on CPAP at 10cmH_2_0 as recommended by UK guidelines(22) and the initial study from Milan(8). Clinical status reassessed with an ABG after one hour on CPAP. Those who had improved on CPAP 10cm H_2_0 were placed on continuous CPAP for 72h as recommended by an Italian protocol(23) with breaks for eating and drinking. Those who failed to improve sufficiently on 10cmH_2_0 of CPAP were given a trial of 15 cmH_2_0 of CPAP(24). If patients improved significantly on 15cmH_2_0, that pressure was continued. Otherwise 10cmH_2_0 was used for all patients given the risks of pressures >10cm H_2_0 such pneumothorax or pneumomediastinum. All patients on oxygen including those on CPAP were encouraged to do prone positioning(25) for at least 1 hour three times daily(26). Those unable to tolerate complete proning were encouraged to do semi prone positioning.

**Figure 1.**
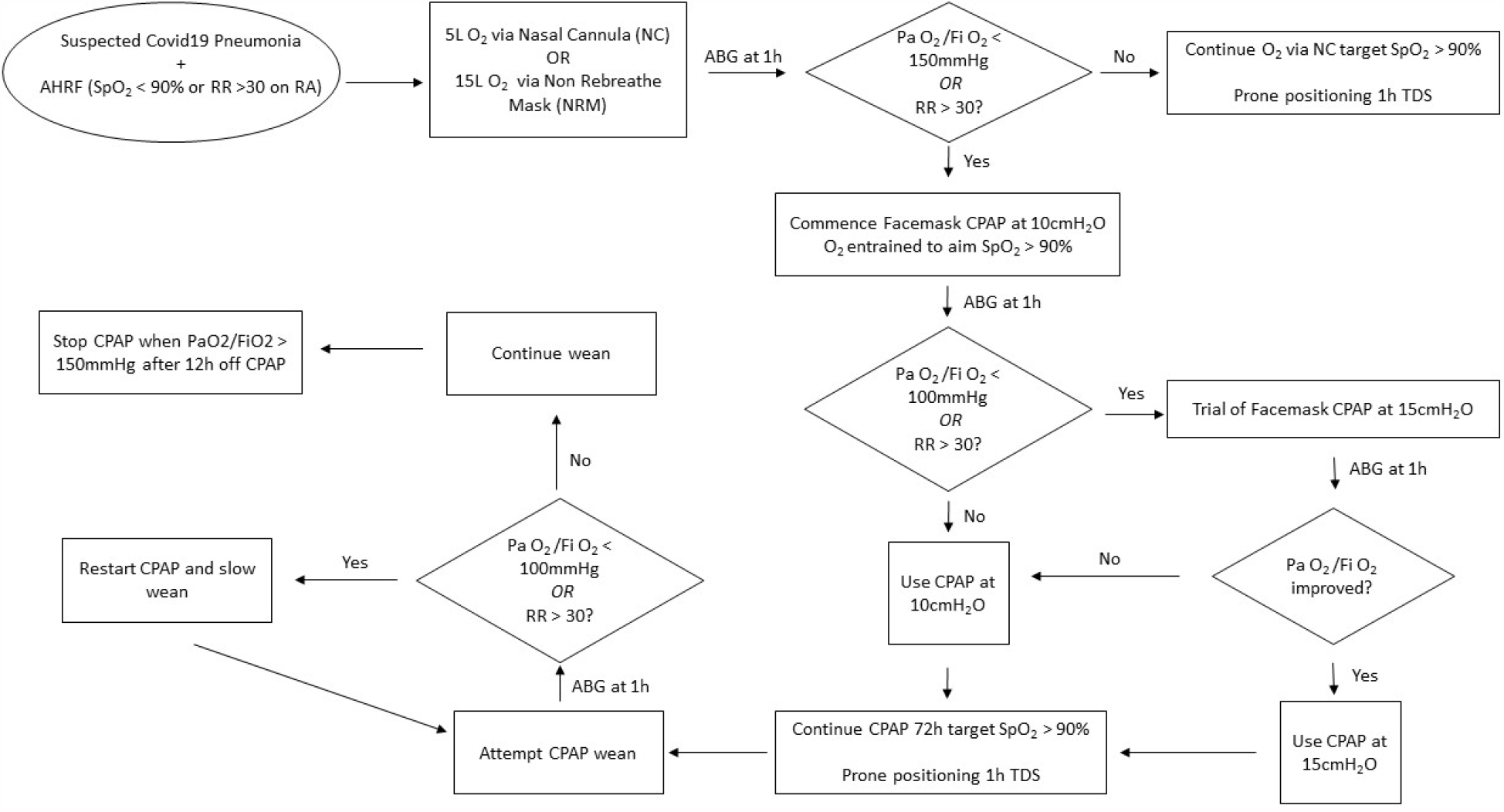
Respiratory Support Algorithm for Covid19 AHRF.

#### CPAP weaning

CPAP weaning was attempted after 72h of continuous CPAP with an ABG after 1h off CPAP to check tolerance of the wean. The rate of the wean was guided by respiratory status off CPAP determined by clinical status and ABGs. The weaning pattern typically consisted of continuous CPAP being followed by a few hours of CPAP in the morning and evening and overnight, then evening and overnight and finally just overnight. CPAP was stopped when evening P_a_O_2_/FiO_2_ was greater than 150mmHg or maintaining SpO2 > 90% on 5L O2 via NC. Patients who failed to wean off CPAP at all were given another 72h of continuous CPAP and a wean was reattempted afterwards.

### Other Medical Management

Medical management of the patients with Covid19 and AHRF is summarised in table 2. Steroids with appropriate thromboprophylaxis formed the mainstay of management. Remdesivir was not used due to lack of evidence of efficacy in severe disease(28), and Tocilizumab was not used due to cost restraints and limited evidence base(32).

**Table 2.** Medical Management for Covid19 AHRF

| Medication | Dosing | Rationale |
| --- | --- | --- |
| Dexamethasone | 1. 6mg OD IV – All patients<br>2. 20mg OD IV for 5 days and then 10mg OD IV for 5 days – Patients with severe ARDS (PaO<sub>2</sub>/FiO<sub>2</sub> < 100mmHg) | 1. Low dose Dexamethasone for all patients as widely advocated by the WHO and others based on multiple studies(27,28).<br>2. Higher dose Dexamethasone based on limited evidence(29) |
| Rivaroxaban | 1. 10mg HS PO – All patients<br>2. 15mg BD PO – Suspected Pulmonary Embolus (PE)(30) | 1. Thromboprophylaxis(28) in the limited local availability of s/c Low Molecular Weight Heparin (LMWH)<br>2. Venous Thrombo-embolism (VTE) Treatment dose for PE |
| Ceftriaxone + Azithromycin | Ceftriaxone 2G OD IV for 10 days + Azithromycin 500mg OD PO for 7 days<br>- All patients | Empirical cover for bacterial superinfection(24) in the context of limited availability of sputum/blood culture facilities |
| Esomeprazole | 20mg OD PO – All patients | Gastroprotection and stress ulcer prophylaxis |
| Ivermectin | 6mg BD PO (4 doses) – All patients | *Strongyloides* prophylaxis in the context of steroid use(31) |

### Other monitoring

All patients had CBC and CXR on admission to support diagnosis of Covid19 Pneumonia. Creatinine and electrolytes for AKI, ECG and RBS to reveal any underlying cardiac disease or diabetes. Blood sugars were monitored daily for steroid induced hyperglycaemia, and strict input/output monitoring was kept to guide a conservative fluid management strategy in the setting of ARDS.

## Results

### Primary Outcome: CPAP Survival Rate for Covid19 Moderate to Severe ARDS

61 patients were admitted with suspected Covid19 AHRF at BCH from April 9^th^ to May 31^st^ 2021 (fig3). 19 patients recovered without requiring escalation to CPAP, while 1 patient died in hospital from suspected massive Pulmonary Embolism (sudden onset tachycardia leading to hypotension and shock, with unsuccessful thrombolysis). 41 patients met criteria for moderate to severe ARDS and were commenced on CPAP regardless of age or underlying comorbidities. 4 patients left against medical advice (LAMA) after being commenced on CPAP: 1 male due to failure to improve on CPAP at 10 days (died), 1 male midway through successful CPAP wean due to financial anxiety despite reassurance (died), 2 females after successful weaning off CPAP due to death of their husbands (both survived). 37 patients were treated with CPAP as per protocol of which 30 patients survived and were discharged while 7 died during hospital admission. Therefore, the survival rate for CPAP patients including the LAMA group was 78% (32/41) overall, while the survival rate among those treated as per protocol was 81% (30/37). The survival rate for all patients admitted to the unit in this period was 83.6% (51/61).

**Figure 3.**
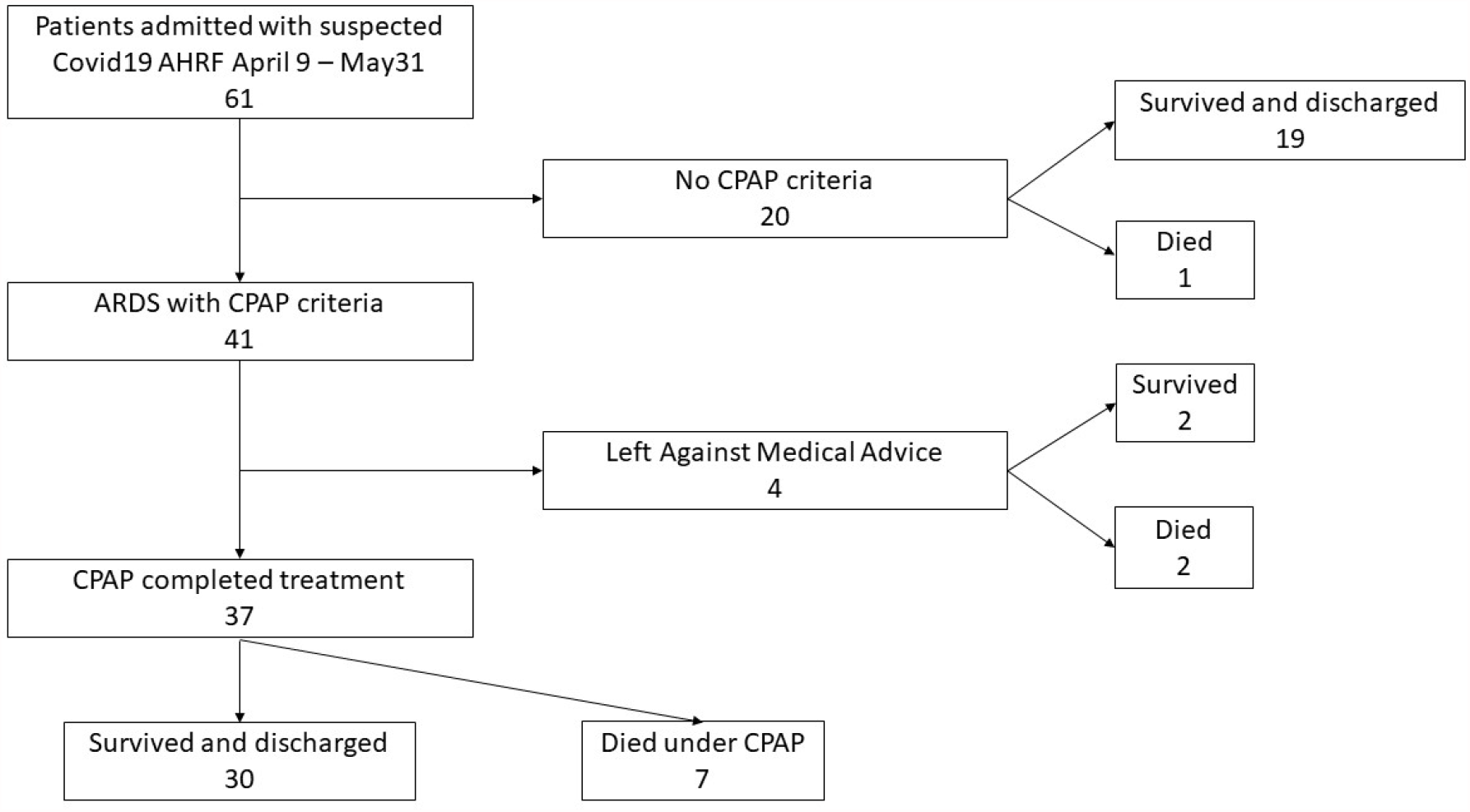
Outcomes of patients admitted with Covid19 AHRF between April 9th and May 31st, 2021.

6 of the 7 patients who died on CPAP died within 2 weeks of admission, of which 1 had developed pneumomediastinum and 1 had pneumothorax and pneumomediastinum– both of which are recognised complications of Covid19 patients on CPAP(33). Chest tube drainage of the pneumothorax was unsuccessful. The 7^th^ patient died after more than 1 month of admission due to suspected bacterial superinfection in the form of hospital acquired pneumonia. Most patients tolerated CPAP well, however some needed mild sedation in the initial stages to improve compliance.

### Secondary outcome 1: Improvement of oxygenation on CPAP

CPAP at 10cmH_2_0 was associated with a significant improvement in oxygenation among moderate to severe ARDS patients with a mean percentage increase of P_a_O_2_/FiO_2_ of 37.2% (*95% CI 17.3 % - 57.1%, n = 31 due to incomplete ABG data*) (fig 4). On initiation of CPAP, patients with severe ARDS (P_a_O_2_/FiO_2_ < 100mmHg) had a greater mean percentage increase in P_a_O_2_/FiO_2_ (*mean increase = 45.8%, 95% CI 18.1 - 73.5%, n =21*) than patients with moderate ARDS (P_a_O_2_/FiO_2_ = 100 – 150mmHg) (*mean increase 19.1 %, 95% CI 2.3 – 35.9 %, n = 10*), but this difference was not statistically significant (p > 0.05).

**Figure 4.**
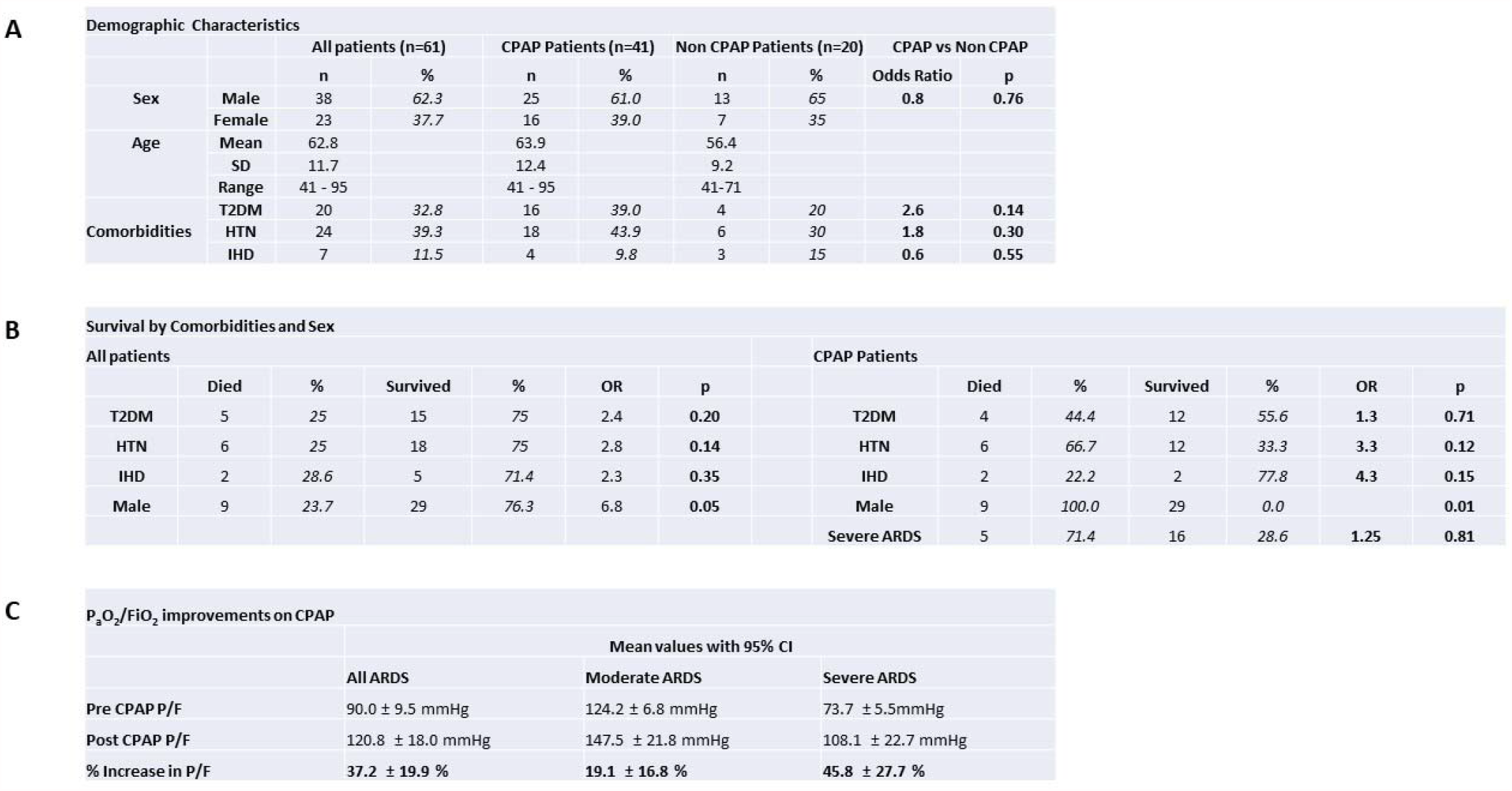
(A) Demographic characteristics of all patients, those requiring CPAP and those who did not require CPAP. (B) Survival of patients by Co-morbidities and Sex. (C) Improvements of P_a_O_2_/FiO_2_ of all patients on CPAP, subdivided into Moderate (P_a_O_2_/FiO_2_ = 100 – 150mmHg), and Severe (P_a_O_2_/FiO_2_ < 100mmHg) ARDS. OR = Odds Ratio, CI = Confidence Interval, P/F = P_a_O_2_/FiO_2_, T2DM = Type 2 Diabetes Mellitus, HTN = Hypertension, IHD = Ischaemic Heart Disease.

### Secondary outcome 2: Predictors of survival – Sex, Comorbidities and Age

Male gender was associated with significant greater mortality overall and on CPAP (p < 0.05). Type 2 Diabetes (Odds Ratio (OR) = 2.4), Hypertension (OR = 2.8) and Ischaemic Heart Disease (OR = 2.3) were associated with increased mortality overall, and this was mirrored in mortality on CPAP. However the differences were not statistically significant (p > 0.05). Age was also not associated with statistically different outcomes in mortality (*Mean age with 95% CI: Patients that survived = 61.6 ± 3.3 years, Patients that died = 62.3 ± 7.1 years; p > 0.05*).

## Discussion

### Primary outcome: Survival on CPAP

Multiple studies have been conducted of patients managed on CPAP for CARDS, but to our knowledge this is the largest case series to date of patients managed exclusively on CPAP. Additionally, this study is also significant coming from a resource poor setting which now accounts for the majority of new Covid19 cases. Our survival rate on CPAP (81% of patients treated as per protocol, 78% of total patients on CPAP) is slightly lower than the 83% reported in the original study on CPAP in CARDS from Milan(8). However, the original study excluded a significant number of patients who were not considered fit for resuscitation, while all patients received CPAP as per protocol in this study regardless of pre-morbid state. Our survival rate is significantly higher than the 29% reported on patients exclusively treated with CPAP in another Italian series(34), however it is likely that these patients had a greater frequency of underlying comorbidities as they were classed ineligible for intubation. The CARDS survival rate for CPAP at our centre is similar to reported rates (around 80%) from Intensive Care Units employing NIV and IMV in resource rich settings(17), implying non-inferiority as a treatment modality. Even more significantly, the CARDS survival rate is significantly higher than that reported from Intensive Care Units in resource poor settings(6,7). With the added benefit of CPAP being significantly less resource intensive in terms of staffing and equipment, and the advantage of health workers lacking critical care background being able to be trained rapidly in its utility, CPAP is an ideal treatment modality for CARDS in the context of the Covid19 pandemic surge in resource limited settings.

Earlier arguments favouring the use of IMV over CPAP within the context of CARDS largely focussed on lack of evidence in its efficacy and concerns around health worker infection from aerosolization during CPAP(2,19). However, this observational study is part of a growing body of evidence for the efficacy of CPAP, and no symptomatic Covid19 infections were recorded among our staff during the period of this study. This was despite SARS COV 2 vaccination levels among staff being low as the vaccination programme was only just being rolled out. Remaining concerns regarding CPAP include complications such as pneumothorax and pneumomediastinum which are associated with higher CPAP pressures (>10cm H_2_0)(33) and occurred in two of our patients with poor outcomes. However, the frequency of these complications is not known to be significantly more with CPAP than with IMV which would have been the alternative in these cases(35). Finally, some studies suggest that the efficacy of CPAP in CARDS is due to a subsection of patients having underlying co-morbidities of CHF and COPD which are well established indications for CPAP. However, this is unlikely to be the case in this study as many patients did not have any premorbid conditions and yet demonstrated improvement on CPAP. Even so, more research is needed to identify which subsection of patients benefitted most from CPAP.

### Secondary outcomes: Improvement in oxygenation on CPAP and predictors of survival

The secondary outcome of the degree of improvement of P_a_O_2_/FiO_2_ (37.2%) was similar to a previous large multicentre study in Northern Italy(45.3%)(36). Improvements in PaO2/FiO2 were greater in the severe ARDS group compared with moderate ARDS but the difference was not statistically significant likely due to sample size. Similarly male gender and co-morbidities of diabetes, hypertension and ischaemic heart disease are known to be predictors of poor outcomes, as reflected in this study. Again however, the lack of statistically significant differences in categories other than gender is likely due to small sample size. Increased age is also known to be a predictor of poor outcome, but the lack of statistically significant difference in our study is likely due to small sample size.

### Limitations

Significant limitations of this study include its observational nature, retrospective single centre design and small sample size. Further research needs to be done to determine the optimal protocol for treatment of CARDS on CPAP, including patient selection, timing of initiation, pressures used and weaning protocols.

## Conclusion

CPAP is an efficacious and cost-effective modality of treatment for Covid19 ARDS (CARDS), particularly in resource poor settings. Survival rates can be demonstrated non inferior to settings employing IMV in resource rich settings. CPAP is less resource intensive in terms of equipment and staffing, and health workers can be easily trained in its operation. CPAP should strongly be considered and promoted for combatting CARDS within context of the pandemic surge of Covid19 now largely occurring in resource poor settings.

## Data Availability

Please contact corresponding author via email

## Acknowledgements

Many thanks to our dedicated nursing staff for their work with Covid19 patients and help with data collection, particularly Nurse in Charge Rizwan Hameed and staff nurse Shahzad Gill. We are thankful to our previous Medical Superintendent Dr Luke Cuthrell, current Medical Superintendent Dr Musheer Shaukat, Hospital Administrator Mr Zubaid Inayat and Nursing Superintendent Nabila Michael for their support and encouragement with the work of our Covid19 unit.

## Conflicts of interest and funding

Authors declare no conflict of interest. No funding was received for this research.

## Notes

### Competing Interest Statement

The authors have declared no competing interest.

### Funding Statement

No funding received

### Author Declarations

Bach Christian Hospital Ethics Committee. Exemption ID: BCH2021/01

